# Children have similar RT-PCR cycle threshold for SARS-CoV-2 in comparison with adults

**DOI:** 10.1101/2021.04.20.21255059

**Authors:** Márcia Polese-Bonatto, Ivaine Tais Sauthier Sartor, Fernanda Hammes Varela, Gabriela Luchiari Tumioto Gianinni, Thaís Raupp Azevedo, Luciane Beatriz Kern, Ingrid Rodrigues Fernandes, Gabriela Oliveira Zavaglia, Caroline Nespolo de David, Walquiria Aparecida Ferreira de Almeida, Victor Bertollo Gomes Porto, Marcelo Comerlato Scotta, Renato T. Stein, COVIDa study group

## Abstract

**Background:** The viral dynamics and the role of children in the spread of SARS-CoV-2 are not completely understood. Our aim was to evaluate RT-PCR Ct values among children with confirmed SARS-CoV-2 compared with that of adult subjects.

**Methods:** Patients (aged from 2 months to ≤18 years, and adults) with signs and symptoms of acute SARS-CoV-2 infection for less than 7 days, were prospectively enrolled in the study from May to November 2020. All participants performed RT-PCR assay for SARS-CoV-2 detection; Ct values of ORF1ab, N, and S gene-targets, and the average of all the three probes were used as surrogates of viral load.

**Results:** There were 21 infants, 62 children and 293 adults of 376 participants with confirmed SARS-CoV-2 infections. RT-PCR Ct values of children under 18 were not significantly different from that of adults after adjusting for days of illness, as observed by the analyzed probes (namely ORF1ab, N, and S), and by the mean of all 3 gene-targets.

**Conclusions:** Ct values for children were comparable to that of adults. Days of illness are a major confounder for SARS-CoV-2 viral load and must be adjusted in any comparison. Although viral load is not the only determinant of SARS-CoV-2 transmission, children may play a significant role in the spread of in the community.

## Introduction

In December 2019, an outbreak of a new viral pneumonia was identified in Wuhan, China^1^. Phylogenetic analysis of the new virus, classified initially as 2019 nCoV, pointed it as a member of the *Betacoronavirus* genus. This virus presented typical features of the coronavirus family previously identified in humans, bats and other wild animals. By February 2020, the Coronavirus Study Group (CSG) of the International Committee on Virus Taxonomy suggested a nomenclature change to severe acute respiratory syndrome coronavirus 2 (SARS-CoV-2) due to phylogenetic analysis relating this new virus to coronaviruses^2^.

All countries have been dealing with this pandemic with an unprecedented number of severe and also non-severe patients. Still, children have somehow been spared from these severe outcomes. An important issue that still needs clarification relates to the understanding of the impact of viral load at different levels of disease, since these could be associated not only with disease severity, but also to its impact on infectivity and viral transmission in the community. The difference on viral load levels between infected children and adults is at this stage of knowledge yet a matter of some debate^3,4^. A study in the US of 145 subjects including children younger than age 5 years, 5 to 17, and adults with mild to moderate illness has found that there was no difference in RT-PCR cycle threshold (Ct) values between older children and adults. Interestingly children younger than age 5 presenting even lower Ct levels than the other age groups^3^. It is important to note that this analysis did not take into account time of symptom onset to the actual testing of the subjects, a possible confounder that also needs to be addressed. An even larger sample size study in Switzerland has shown similar results^5^.

The viral load of SARS-CoV-2 can be indirectly measured using the Ct value of automated RT-PCR techniques, and its value is inversely proportional to the viral load. Ct value is defined as the number of cycles necessary to amplify viral RNA to reach a detectable level, which is a positive fluorescent amplification signal in a real-time reverse transcription-polymerase chain reaction (RT-PCR) assay^6,7^. Pujadas et. al.^8^ suggested the use of viral load to identify severe patients, and to develop predictive algorithms.

The role of children as spreaders of SARS-CoV-2 is not completely understood, even though it is clear that they have been spared of severe presentations of the disease^3,9^. However, such differences among different age groups remain unclear^10^. In this study we report the viral RNA Ct values patterns observed during the early phase of infection in a cohort of patients with confirmed SARS-CoV-2 infection and have these Ct values compared between children and adults, also taking into account time from symptom’s onset to PCR testing.

## Materials and Methods

This was a prospective cross-sectional multicenter study with data collected in two hospitals in Brazil. From May to November 2020, we assessed outpatient subjects seen at these emergency rooms (ERs), or those hospitalized, presenting with at least one sign or symptom suggestive of COVID-19 (cough, fever, or sore throat) within 7 days of symptoms onset (SO). Eligible patients with confirmed SARS-CoV-2 infection were included in the study. Age groups compared were classified as children (aged from ≥2 mo to <18 years) and adults (≥18 years). In order to explore possible differences among childhood age groups, children were further subgrouped as infants (2 mo. to <2 years) and older children (aged from ≥2 years to <18 years).

All participants performed RT-PCR assay for SARS-CoV-2 detection. Procedure for sample collection involved one bilateral nasopharyngeal and one oropharyngeal swab collection. Both swabs were placed in the same transport medium with saline solution and RNAlater®, RNA Stabilization Solution (Catalog number AM7021, Life Technologies, Carlsbad, CA, USA). RNA was extracted using MagMax™ Viral/Pathogenic Nucleic Acid Isolation (Catalog number A48310, Applied Biosystems, Austin, Texas, EUA) in the KingFisher Duo Prime System platform (ThermoFisher Scientific, Waltham, Massachusetts, USA). The RT-PCR assay was and performed using Path™ 1-Step RT-qPCR Master Mix, CG (catalog number A15299, AppliedBiosystems, Frederick, Maryland, USA) and TaqMan™ 2019-nCoV Assay Kit v1 (catalog number A47532, ThermoFisher Scientific, Pleasanton, California, EUA) in 10 µL total reaction, of which 5 µL were RNA. For reaction control we used 5 µL (200 copies/µL) of TaqMan™ 2019-nCoV Control Kit v1 (catalog number A47533, ThermoFisher Scientific, Pleasanton, California, EUA). Due to assistance and laboratory decisions, since 7^th^ October, the RT-PCR assays were performed using only for *N* and *ORF1ab* SARS-CoV-2–specific targets.

We have excluded participants with clinical conditions that compromised the immune system that could potentially interfere with viral load, such as type 1 or 2 diabetes mellitus, previous organ transplant, cancer diagnosis or subjects who had chemotherapy in the two weeks previous to enrollment.

We included in these analyses only participants within the first 7 days of SO, since the peak of infectivity largely occurs in the first week of illness^11^. All the three probes Ct values of *ORF1ab, N, S* gene-targets and the median of these were compared as surrogates of viral load. Percentages were used to describe categorical variables; continuous variables were summarized in terms of median and interquartile range (IQR). Data normality assumptions were verified for continuous variables. Two-tailed Mann-Whitney-Wilcoxon test or two-tailed Kruskal-Wallis test followed by Benjamini-Hochberg correction for multiple comparisons, used to compare Ct values of *ORF1ab, S* and *N* SARS-CoV-2– specific targets between groups. Linear regression analysis was performed to associate the Ct values from RT-PCR assays with age group and days of SO, and odds ratio with 95% confidence intervals were calculated. The main analysis was the comparison of Ct values between children and adults. Secondary analysis was the comparison among 3 age groups (infants, older children and adults), as well as between infants versus older children and adults. All data preprocessing and analyses were performed in R 3.5.0 statistical software^12^.

The study was performed in accordance with the declaration of Helsinki and Good Clinical Practice Guidelines, after approval by the Hospital Moinhos de Vento Institutional Review Board (IRB number 30749720.4.1001.5330) submitted April 14th, 2020 and a decision made April 17th, 2020 (decision number 3.977.144) with approval. All participants included in this study provided either a written informed consent, or a legal responsible provided written informed consent.

## Results

In the study 1,823 subjects were originally screened, 1,113 were excluded due to a negative SARS-CoV-2 RT-PCR result, and 334 excluded for other reasons, as shown in Figure 1. There were 376 participants with confirmed SARS-CoV-2 infection, stratified as infants (N = 21, 5.6%), older children (N = 62, 16.5%), and adults (N = 293, 77.9%). 10.6% of the adults (31/293) required hospitalization, while almost all infants and children had only mild clinical symptoms with only one child needing admission but not due to Covid, and not progressing for a severe outcome. All infants were outpatients, whereas among children and adults, 324 (91.3%) participants were seen as outpatients; 28 (7.9%) were admitted (including one child): 25 to infirmary and 3 (0.8%) to ICU (all adults), respectively.

**Figure 1.**
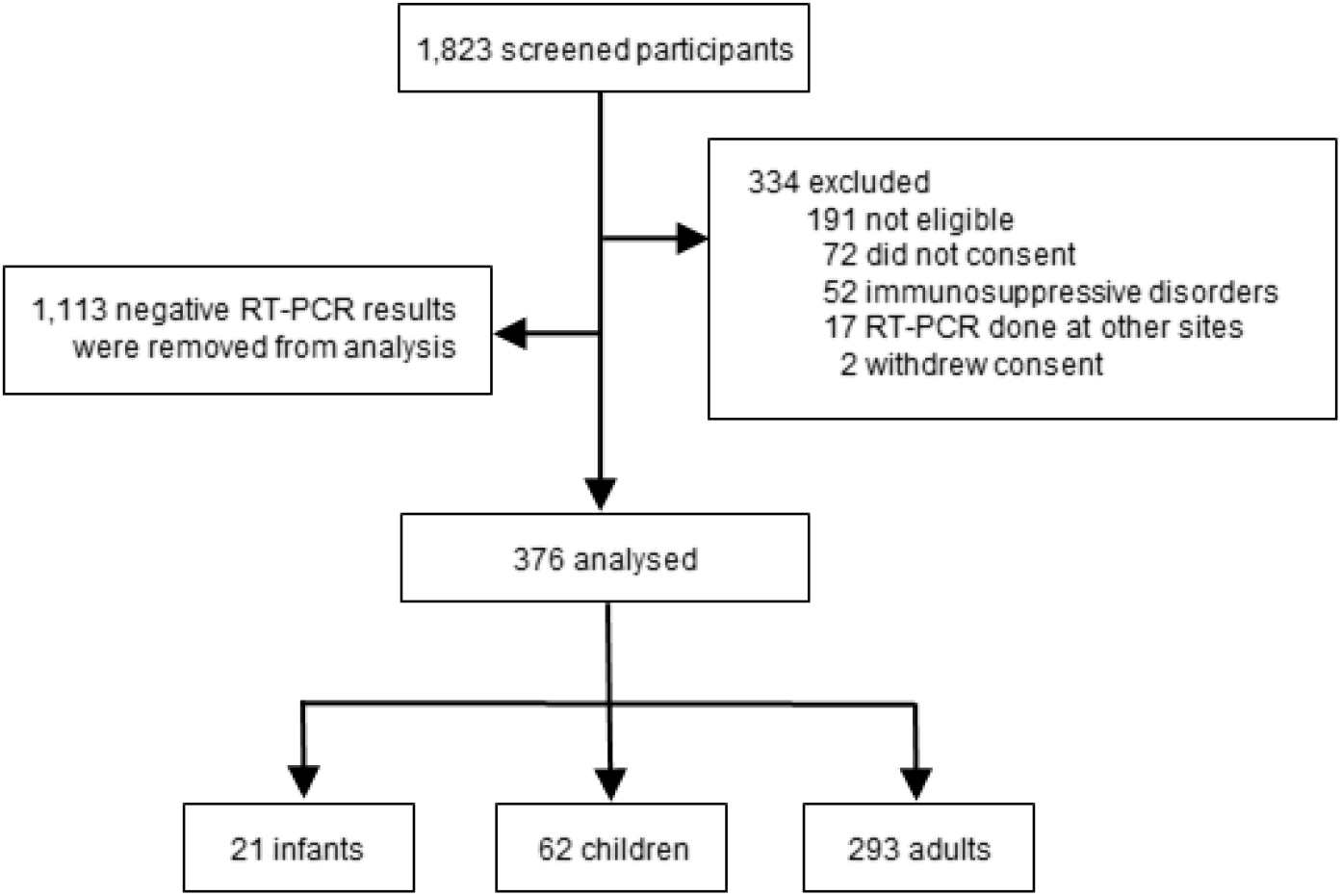
Study subjects’ flowchart.

For the initial analysis participants were split in two groups, of <18 years and ≥18 years of age, to compare overall children and adult mean Ct values. Mean Ct values for children (<18 years) and adults were 20.73 and 20.83 (P = 0.453), respectively, for all gene targets (Figure 2). When children were further stratified in two age groups (2 months to <2 years, and ≥ 2 to 18 years), mean Ct values were 18.11 for infants, 21.19 for children, and 20.84 for adults (P = 0.128) for all gene targets, as shown in Figure 3. In a subsequent analysis, older children and adults were then combined in a single group, and compared to infants. Median Ct values for *ORF1ab, S*, and for the median of all the three gene-targets were significantly lower for infants group P = 0.037, P = 0.006, P = 0.044, respectively) (Figure 4).

**Figure 2.**
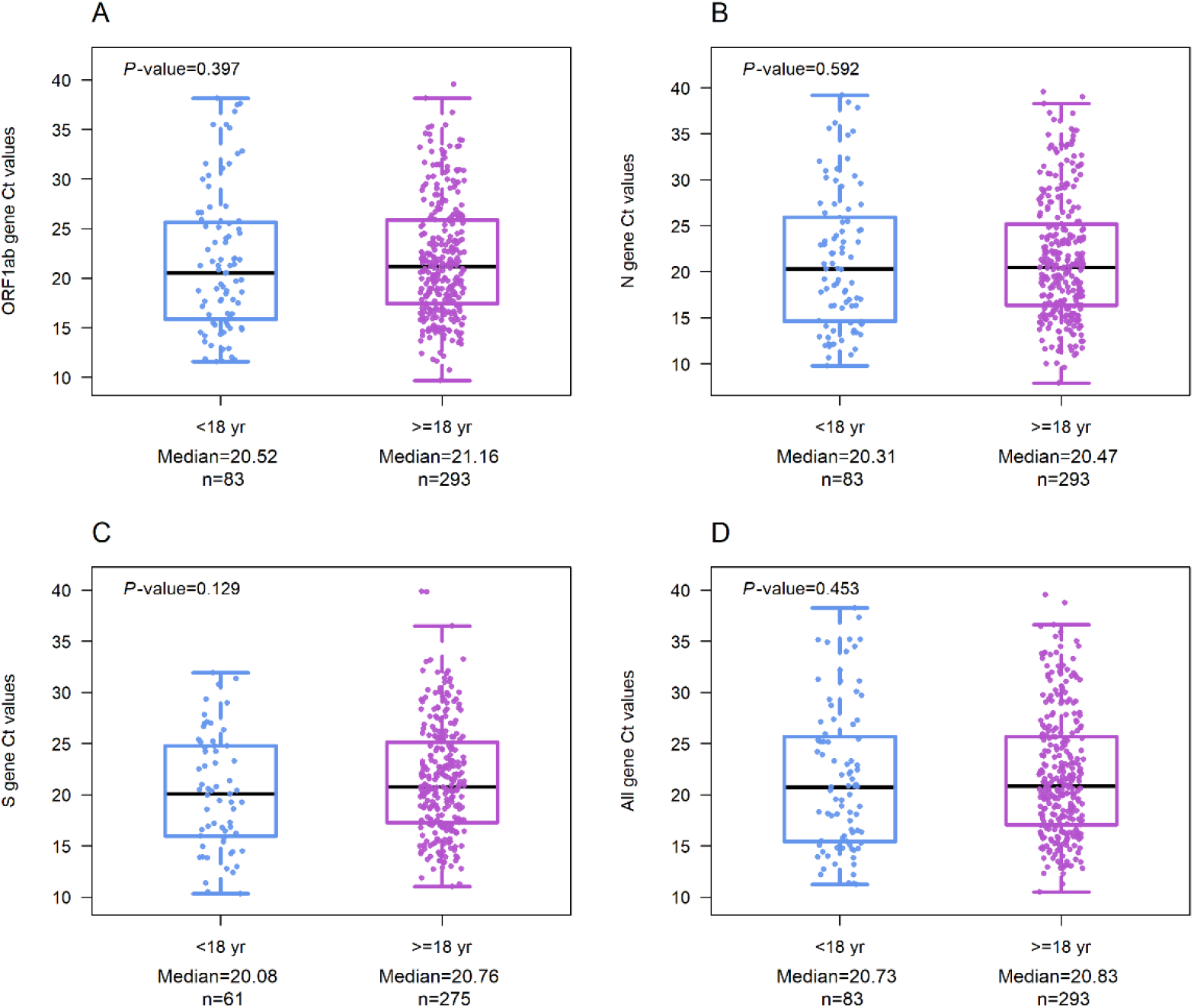
Comparison of Ct values in those of 2 months to 18 years versus adults with confirmed SARS-CoV-2 infection. (A) Ct values of *ORF1ab* gene. (B) Ct values of *N* gene. (C) Ct values of *S* gene. (D) Ct values of the mean of *ORF1ab, N* and *S* genes. Median is represented as a solid black line, interquartile ranges are represented by boxes, upper and lower adjacent values are represented by whiskers, and outliers are represented by isolated points. (mo) months. (yr) years.

**Figure 3.**
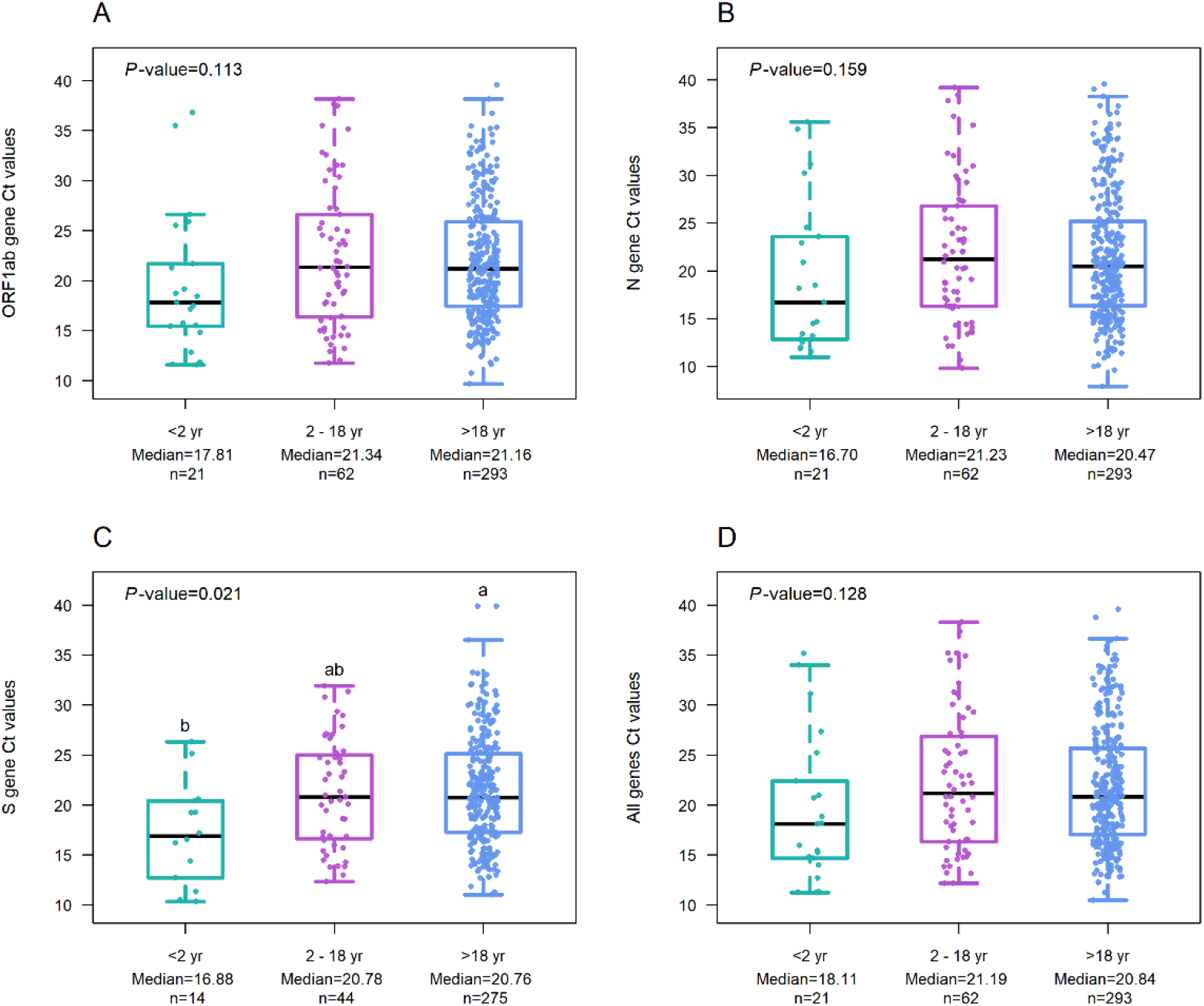
Comparison of Ct values according to age stratification among participants with confirmed SARS-CoV-2 infection. (A) Ct values of *ORF1ab* gene. (B) Ct values of *N* gene. (C) Ct values of *S* gene. (D) Ct values of the mean of *ORF1ab, N* and *S* genes. Median is represented as a solid black line, interquartile ranges are represented by boxes, upper and lower adjacent values are represented by whiskers, and outliers are represented by isolated points. (mo) months. (yr) years.

**Figure 4.**
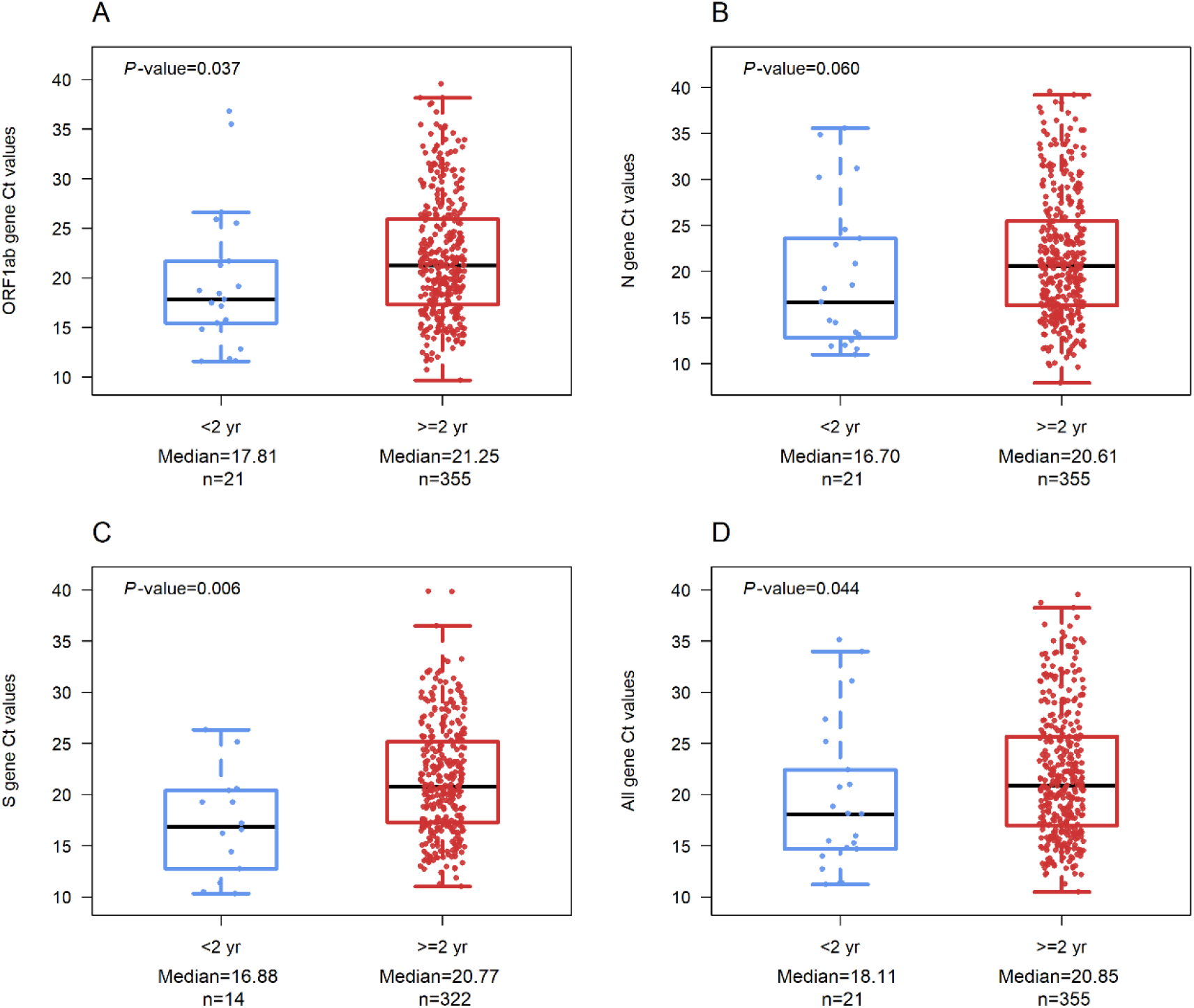
Comparison of Ct values of infants *versus* children and adults with confirmed SARS-CoV-2 infection. (A) Ct values of *ORF1ab* gene. (B) Ct values of *N* gene. (C) Ct values of *S* gene. (D) Ct values of the mean of *ORF1ab, N* and *S* genes. Median is represented as a solid black line, interquartile ranges are represented by boxes, upper and lower adjacent values are represented by whiskers, and outliers are represented by isolated points. (mo) months. (yr) years.

Median of SO to the time of sample collection was lower in infants (2 days, IQR 1-3, range 1-7) than for those ≥2 years (3 days, IQR 2-4, range 0-7) with *P* = 0.01. Additionally, when Ct values of infants compared to adults considering days of SO were no longer different (OR = 5.52 (0.37-83.24), P = 0.217), indicating that infants have equal Ct values than adults (Supplementary Table 1). Days of SO were statistically significant (OR = 2.09 (1.48-2.93), P < 0.001). Demographic and baseline clinic details are presented in Supplementary Table 2.

## Discussion

Our findings reinforce previous findings suggesting that children do not have lower SARS-CoV-2 viral loads as compared to adults^13^. Despite infants with positive SARS-CoV-2 testing having lower Ct values when compared to other age groups, such difference was not significant after adjusting for days of SO, since infants were enrolled earlier in the course of disease. Our findings clearly suggest that days of SO are a major factor in SARS-CoV-2 viral load and any analysis must be adjusted for this feature. This is important because some previous publications have not accounted for this important confounding variable. It is important to remember that there was no clinical correlation with these low Ct leves (thus, high viral load) in infants, since they all presented very mild disease.

As suggested by Yonker et al^4^ children could carry higher viral loads even with mild symptoms when a sample is obtained in an early infection phase. Other study also point out to the fact that children younger than five years of age presented higher viral load when compared with older children and adults^3^. However, recent viral load comparisons across age groups have yielded inconsistent conclusions^5,13^. Some studies following adults only show a positive correlation between higher SARS-CoV-2 viral load with disease severity, need of mechanical ventilation, and/or higher mortality rates, which is not the case in children^14^. The reasons for such differences in the relation of viral load and severity across age groups are not yet fully understood^14^, but could well be related to different testing times in relation to symptom’s onset.

Although viral load may not be the only determinant of transmission of SARS-CoV-2, it is probably one of the most important factors. On the other hand, as children usually present with a milder form of disease, the lower frequency of cough and the generation of droplets and aerosols may have some impact in reducing the risk of aerosolized transmission. However, despite the usual milder disease in children, our findings highlight that efforts to mitigate transmission should include this age group, since we did not find any difference in Ct levels between overall children and adults. The finding of (at least) similar viral load in infants is of some concern, since most current guidelines do not recommend that this age group should wear face masks in public places^15,16^.

Our study has some limitations worth mentioning. We have included only symptomatic participants during the early stage of SARS-CoV-2 infection, meaning we cannot draw conclusions about asymptomatic individuals or those presenting later in the course of the disease. Another limitation was a restricted sample size of infected children. Social distancing, closed schools and daycare centers, together with the concern to take children to ERs have led to a sharp reduction in the overall number of pediatric consultations. Therefore, our sample could be underpowered to detect greater differences between age groups. It also raises an important issue of undiagnosed pauci-symptomatic or asymptomatic children spreading the virus in the community^5,17,18^. Also, the quality and volume of viral RNA on collected swabs could have varied depending on how the sample collection was conducted^19^. We have not corrected our sample for the amount of viral RNA, since it would only add another step to the diagnostic routine, which was already overstretched at that point of the pandemic. But it is well known that cycle threshold values can be affected by a batch effect^20^, since variations among different runs can occur.

Despite these limitations our findings are strong enough to suggest that symptomatic children may play a significant role in SARS-CoV-2 transmission since they harbor viral loads that are not lower than adults. Our findings suggest that in earlier stages of disease there is greater chance of higher viral load and possible higher risk of transmission. Our data indicates that the return of school for older children and daycare activities for the younger ones must be made with caution due to the impact in spreading the infection by these groups, who do not yet have a vaccination perspective.

Our findings strongly suggest that symptomatic children have equivalent Ct values when compared to adults, and even more interestingly higher viral loads seem to be great very early in the disease, even in the presence of mild clinical presentation.

## Data Availability

Authors declare that al data is available under request.

## Acknowledgments

We thank the Scientific Committee of the Research Support Nucleus (NAP) of Moinhos de Vento Hospital for technical-scientific consultancy. We thank the statistician Matias Segelis Vieira who supported data analysis. We thank the inclusion personal, laboratory team, and site staff from Hospital Moinhos de Vento and from Hospital Restinga e Extremo Sul. Aline Andrea da Cunha, Joao Ronaldo Mafalda Krauser, Paulo Sergio Kroeff Schmitz, Sidiclei Machado Carvalho, Fabio Jose Rockenbach, Kelly Viegas Antunes, Marcelo da Silva Ferreira, Rafael Garcia Trindade, Thayna Silva Lino, Claudia Josiel Oliveira do Coito, Thaís Pacheco, Paloma Bortolini Martins, Bruna de Oliveira Rocha, Darléia Radaelli, Tiago da Silva Silvano, Adriana da Silva Silveira, Alceu Kuckoski, Ana Paula da Silva Lopes, Andreia Escobar da Costa, Clarice Cardoso Machado, Erica Vieira da Silva, Evelin Inácia da Silva, Luciana Rodrigues Ribeiro, Marcely Mayr da Costa, Morgana Thais Carollo Fernandes and Rafael da Silva Cassafuz.

## COVIDa study group

Adriane Isabel Rohden, Amanda Paz Santos, Ana Carolina Monteiro da Rocha, Ana Paula dos Santos, Camila Bonalume Dall’Aqua, Camila Dietrich, Caroline Cabral Robinson, Catia Moreira Guterres, Charles Francisco Ferreira, Débora Vacaro Fogazzi, Denise Arakaki-Sanchez, Eliana Marcia da Ros Wendland, Emerson Silveira de Brito, Fernanda Lutz Tolves, Fernando Rovedder Boita, Francieli Fontana Sutile Tardetti Fantinato, Giovana Petracco de Miranda, Gisele Alcina Nader Bastos, Glaucia Fragoso Hohenberger, Jaina da Costa Pereira, Leonardo Araújo Pinto, Madelaine Correa de Oliveira, Maicon Falavigna, Maristênia Machado Araújo, Patricia Bartholomay Oliveira, Raul Correia da Silva, Regis Goulart Rosa, Shirlei Villanova Ribeiro, Tássia Rolim Camargo, Thainá Dias Luft, Thaís Jacobsen Duarte, Thayane Martins Dornelles, Tiago Fazolo and William Jones Dartora.

---

**Supplementary Table 1.** Linear regressions considering age groups and days of SO for the gene-targets.

|  | Age, years (reference: 2 mo to <2 yr) |  | SO, days (continuous) |  |
| --- | --- | --- | --- | --- |
| Gene-target | OR (95%CI) | P-value | OR (95%CI) | P-value |
| ORF1ab | 5.76 (0.40-83.24) | 0.198 | 1.98 (1.42-2.77) | **<0.001** |
| N | 4.47 (0.23-87.23) | 0.322 | 2.26 (1.55-3.28) | **<0.001** |
| S | 44.95 (2.89-698.64) | 0.007 | 2.40 (1.77-3.25) | **<0.001** |
| All gene targets | 5.52 (0.37-83.24) | 0.217 | 2.09 (1.48-2.93) | **<0.001** |

**Supplementary Table 2.**
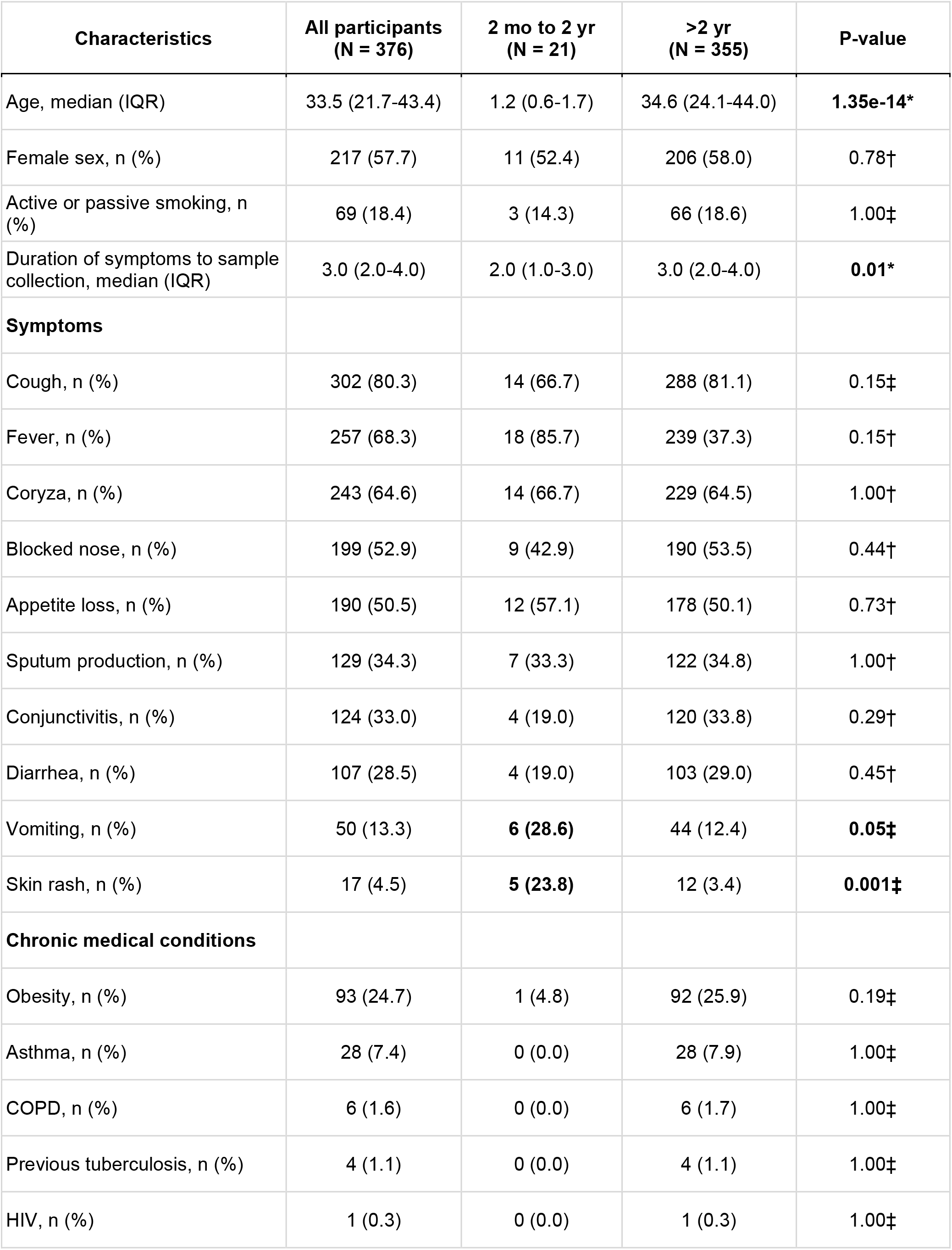
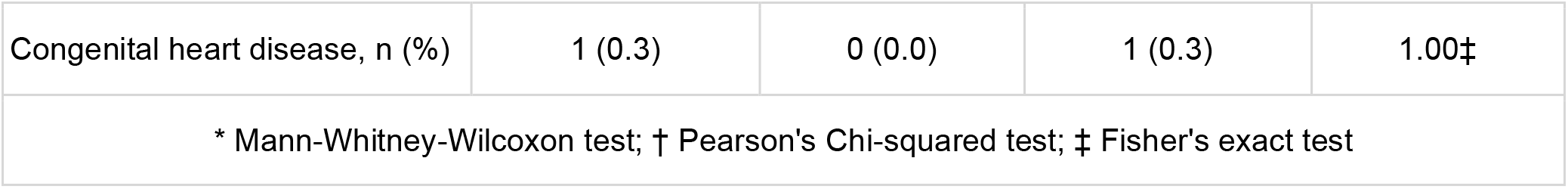
Demographic and clinical characteristics of included subjects.

| Characteristics | All participants<br>(N = 376) | 2 mo to 2 yr<br>(N = 21) | >2 yr<br>(N = 355) | P-value |
| --- | --- | --- | --- | --- |
| Age, median (IQR) | 33.5 (21.7-43.4) | 1.2 (0.6-1.7) | 34.6 (24.1-44.0) | **1.35e-14\*** |
| Female sex, n (%) | 217 (57.7) | 11 (52.4) | 206 (58.0) | 0.78† |
| Active or passive smoking, n (%) | 69 (18.4) | 3 (14.3) | 66 (18.6) | 1.00‡ |
| Duration of symptoms to sample collection, median (IQR) | 3.0 (2.0-4.0) | 2.0 (1.0-3.0) | 3.0 (2.0-4.0) | **0.01\*** |
| **Symptoms** |  |  |  |  |
| Cough, n (%) | 302 (80.3) | 14 (66.7) | 288 (81.1) | 0.15‡ |
| Fever, n (%) | 257 (68.3) | 18 (85.7) | 239 (37.3) | 0.15† |
| Coryza, n (%) | 243 (64.6) | 14 (66.7) | 229 (64.5) | 1.00† |
| Blocked nose, n (%) | 199 (52.9) | 9 (42.9) | 190 (53.5) | 0.44† |
| Appetite loss, n (%) | 190 (50.5) | 12 (57.1) | 178 (50.1) | 0.73† |
| Sputum production, n (%) | 129 (34.3) | 7 (33.3) | 122 (34.8) | 1.00† |
| Conjunctivitis, n (%) | 124 (33.0) | 4 (19.0) | 120 (33.8) | 0.29† |
| Diarrhea, n (%) | 107 (28.5) | 4 (19.0) | 103 (29.0) | 0.45† |
| Vomiting, n (%) | 50 (13.3) | **6 (28.6)** | 44 (12.4) | **0.05‡** |
| Skin rash, n (%) | 17 (4.5) | **5 (23.8)** | 12 (3.4) | **0.001‡** |
| **Chronic medical conditions** |  |  |  |  |
| Obesity, n (%) | 93 (24.7) | 1 (4.8) | 92 (25.9) | 0.19‡ |
| Asthma, n (%) | 28 (7.4) | 0 (0.0) | 28 (7.9) | 1.00‡ |
| COPD, n (%) | 6 (1.6) | 0 (0.0) | 6 (1.7) | 1.00‡ |
| Previous tuberculosis, n (%) | 4 (1.1) | 0 (0.0) | 4 (1.1) | 1.00‡ |
| HIV, n (%) | 1 (0.3) | 0 (0.0) | 1 (0.3) | 1.00‡ |
| Congenital heart disease, n (%) | 1 (0.3) | 0 (0.0) | 1 (0.3) | 1.00‡ |
| * Mann-Whitney-Wilcoxon test; † Pearson's Chi-squared test; ‡ Fisher's exact test |  |  |  |  |

